# The indirect impacts of routine azithromycin prophylaxis during labour on antimicrobial resistance in low-income and middle-income countries: a population-level modelling analysis

**DOI:** 10.1101/2025.09.05.25335177

**Authors:** Ben S Cooper, Aronrag C Meeyai

## Abstract

We aimed to determine the potential population-level impact on antibiotic resistance of a universally adopted policy of azithromycin prophylaxis for pregnant women in labour planning a vaginal birth. We developed three mathematical models with increasing levels of complexity to represent the population processes through which increased use of antibiotics resulting from the policy affects antibiotic resistance. We used each of these models to explore possible impacts of the policy on antibiotic resistance in three important bacterial pathogens: *Streptococcus pneumoniae, Escherichia coli*, and *Staphylococcus aureus*. All three models considered the impacts of the policy on the asymptomatic carriage of macrolide-resistant variants of these three pathogens. We assumed that antibiotic resistance in clinically important infections changes in line with changes in resistance in asymptomatic carriage. The simplest model (model 1) has previously been shown to be able to explain the relationship between antibiotic consumption and antibiotic resistance in *E. coli* and *S. pneumoniae* in European countries, with significant improvements over previously described models for these pathogens. The second model (model 2) extended model 1 to consider interactions between a hospital maternity unit (where azithromycin prophylaxis is delivered) and the wider community. The third model (model 3) extended model 2 by dividing the population into three age groups: infants, children aged 1-12 years, and those aged 13 years and over. All three models found that increased azithromycin usage from adopting the prophylaxis policy would be expected to lead to increased macrolide resistance at a population level in the three pathogens studied, with larger effects found in countries with higher per capita birth rates. Increasing the complexity and realism of the model consistently led to smaller estimates of the impact on resistance. For example, using model 3, it was estimated that the policy would increase macrolide resistance in *S. aureus* and *S. pneumoniae* in Bangladesh by between 8 and 15%. In contrast, increases of 16% to 25% were seen when using model 1. To put these numbers into context, it has been estimated that globally, in 2021, 22,000 deaths were attributable to macrolide resistance in *S. aureus* and 20,000 deaths were attributable to macrolide resistance in *S. pneumoniae*. A 10% increase in macrolide resistance in these pathogens would therefore be expected to lead to an additional 4,200 annual attributable deaths. As with any modelling study there are many caveats: there are many large uncertainties relating to both model structures and parameter values as well as current levels of macrolide use and macrolide resistance.

## Introduction

A single 2-g oral dose of azithromycin administered to women in labour at 28 weeks or more gestation has been shown to reduce a composite endpoint of maternal sepsis or death in an individually randomised trial conducted in six low and middle-income countries in Africa, Asia and Central America (Tita et al. 2023). This prophylactic use of antibiotics was associated with a large reduction in a composite outcome of maternal sepsis or infection in sites in sub-Saharan Africa (relative risk (RR) 0.49, 95% confidence interval (CI) 0.39-0.61); smaller reductions were seen at sites in Asia (RR: 0.79, 95%CI 0.70-0.90) (Carlo et al. 2025). In central America, the number of patients enrolled was too small to draw meaningful conclusions. The trial found no evidence that the intervention reduced neonatal sepsis or death, and there was no evidence of reduced maternal mortality (RR: 1.23; 95% CI, 0.51 to 2.97). Despite these benefits in reducing maternal sepsis and infection, there are concerns that adoption of a policy of universal azithromycin prophylaxis for women in labour could lead to increases in azithromycin resistance in important bacterial pathogens in the wider population. Such an increase in antibiotic resistance could be a cause of worse clinical outcomes in infections caused by these pathogens and, potentially, an increased number of infections. Any direct health benefits of the policy must be considered alongside these potential indirect negative effects. Concerns that universal azithromycin prophylaxis for women in labour will result in increases in azithromycin resistance in clinically important pathogens are supported by evidence from studies showing that oral azithromycin during labour causes a transient increase in the prevalence of bacterial carriage of azithromycin-resistant *S. aureus* in infants of treated mothers and a transient increase in carriage of azithromycin-resistant *S. aureus* and *S. pneumoniae* in treated mothers (Roca et al. 2016). While this elevated resistance in treated individuals wanes over time, reducing to close to baseline levels after 12 months (Bojang et al. 2018), a population-wide adoption of the policy of administering azithromycin to all women during labour would be expected to result in azithromycin-resistance reaching a higher equilibrium value in the wider population. Indeed, substantial long-term increases in azithromycin resistance in *S. pneumoniae* have been observed in communities assigned to the treatment arm in a community-randomised trial of mass drug administration (MDA) of azithromycin to reduce childhood mortality even amongst children in those communities who did not receive the intervention (Kalizang’oma et al. 2025).

The lack of community-randomised trials of azithromycin prophylaxis during labour precludes direct assessment of the population-level impacts of the policy on azithromycin resistance. Instead, in this study we address the question using a suite of dynamic models to explore how the increased use of azithromycin the policy would entail might be expected to affect levels of resistance to azithromycin. We then combine modelling results with previously published estimates of the health burden of azithromycin resistance to quantify the potential adverse consequences of the intervention resulting from such resistance selection.

## Methods

To explore how a policy of administering a single dose of oral azithromycin to women in labour might be expected to affect levels of azithromycin-resistance we implemented a suite of three dynamic models of increasing complexity. The models are run at WHO regional level using country-level estimates of macrolide consumption and macrolide resistance which are weighted by the population sizes of the constituent countries of the WHO regions to obtain regional estimates.

Our baseline model (Model 1) used the deterministic formulation of the “mixed carriage” model described by Davies et al (Davies et al. 2019). In common with other models for drug-resistant bacteria, this model focuses on the asymptomatic carriage of drug-resistant and drug-sensitive bacteria. This is because such asymptomatic transmission drives the spread of the key bacterial pathogens we are concerned with here. This model embeds within-host dynamics within a pathogen transmission model and is structurally neutral in the sense that it meets criteria for both ecological neutrality and population genetic neutrality (Lipsitch et al. 2009). Hosts are grouped into one of five distinct compartments: uncolonized with either antibiotic sensitive or antibiotic resistant strains of the pathogen under consideration (labelled *X* in Figure 1), colonized only with an antibiotic-sensitive strain of the pathogen (*S*), colonized only with an antibiotic-resistant strains of the pathogen (*R*), and colonized with both strains, but with the sensitive strain dominant (*SR*) or with the resistant strain dominant (*RS*). For *S. pneumoniae* and *E. coli* this model has been shown to better capture the relationship between antibiotic use and resistance than simpler models where hosts are exclusively colonised with resistant or susceptible strains, such as that proposed by Olesen 2022 (Olesen 2022). Our analysis also used two extensions of this basic model: first by considering two setting types (community and maternity unit) with low and high rates of antibiotic use (Model 2), and then by extending Model 2 to include four different host types in the community: mothers, infants, other children, and other adults (Model 3).

**Figure 1.**
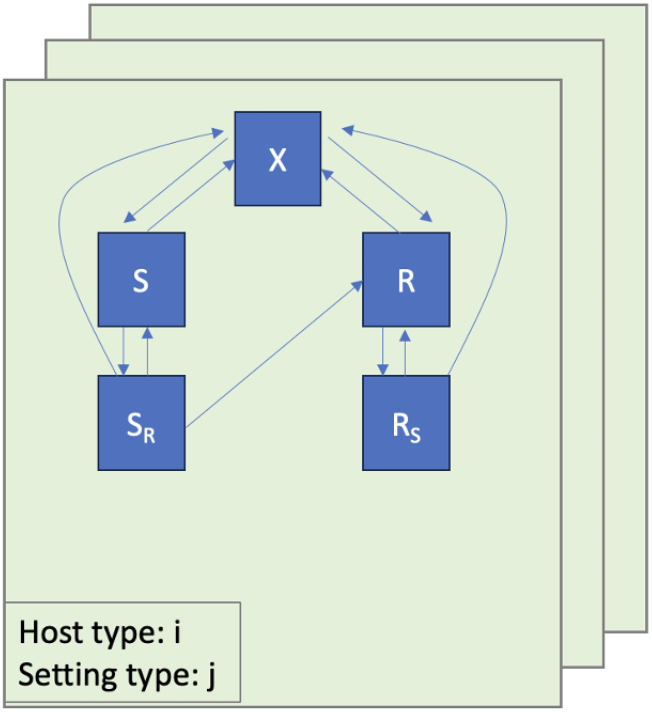
Model structure: In the simplest model (Model 1) there is a single host type and a single setting type. In Model 2 there are two setting types, and in Model 3 there are two setting types and 4 host types. In all models host can be in one of five possible compartments within each setting: not colonised with the focal pathogen (*X*); colonised only with a susceptible strain (*S*); colonized only with a resistant strain (R); colonized with susceptible and resistant strains with susceptible dominant(*SR*); and colonized with susceptible and resistant strains with resistant dominant (*RS*).

In contrast to *E. coli* and *S. pneumoniae*, for *S. aureus* co-colonisation with antibiotic resistant and susceptible strains is much less frequent (Dall’Antonia et al. 2005), and is not needed to explain co-existence of resistant and sensitive phenotypes which can be parsimoniously explained by interactions between settings with high and low levels of antibiotic use, most commonly hospital and community (Kouyos, Klein, and Grenfell 2013). When modelling *S. aureus*, to reflect this greater degree of bacterial interference, we therefore used a value of 0.1 for the model parameter governing the relative efficiency of co-colonisation in contrast to 0.9 for *E. coli* and *S. pneumoniae*.

Baseline values for those model parameters which are assumed not to vary by geographic region are given in Table 1. Model parameters which were assumed to vary by geographic region are the macrolide treatment induced clearance rate (***τ***) and the transmission parameter (β). The baseline macrolide treatment induced clearance rate was based on estimates of the number of completed macrolide courses per person per year. These were derived from estimates of human macrolide consumption in 2018 from Browne et al. (Annie J. Browne et al. 2021) under the assumption that one course is 12 mg/kg for five days. Regional estimates were derived from national estimates by weighting for population size. National UN data from 2020 on the mean number of births per person per year were used to estimate per capita increases in macrolide use that would occur if a policy of universal azithromycin use during labour were to be introduced, and these were again weighted by population size to to produce estimates for each WHO region. For *S. pneumoniae* and *S. aureus* the transmission parameter was estimated for each WHO region using by choosing the value such that, at equilibrium, the predicted proportion of colonised individuals who had a resistant strain dominant (i.e. the R and RS compartments in Figure 1) was equal to estimated proportion of clinical infections with macrolide resistance. For this analysis, the model used 2018 estimates of macrolide consumption (Annie J. Browne et al. 2021) and the estimated proportion of infections with each pathogen in each WHO region that were macrolide-resistant in 2018 based on population-weighted averages of country-level estimates (GBD 2021 Antimicrobial Resistance Collaborators 2024). This analysis therefore implicitly assumes that resistance is at equilibrium and that clinical infections are equally likely to occur in individuals colonised with dominant resistant strains as they are in individuals colonised with dominant susceptible strains. For *E. coli* global estimates of macrolide resistance were not available, and we instead performed a sensitivity analysis where we varied the assumed baseline proportion of *E. coli* infections that were macrolide resistant from 10% to 30% and determined the values of the models’ transmission parameters that would be needed to match these proportions. Full details of model parameters can be found in Appendix 2 and model can be found <insert github link>.

**Table 1.**
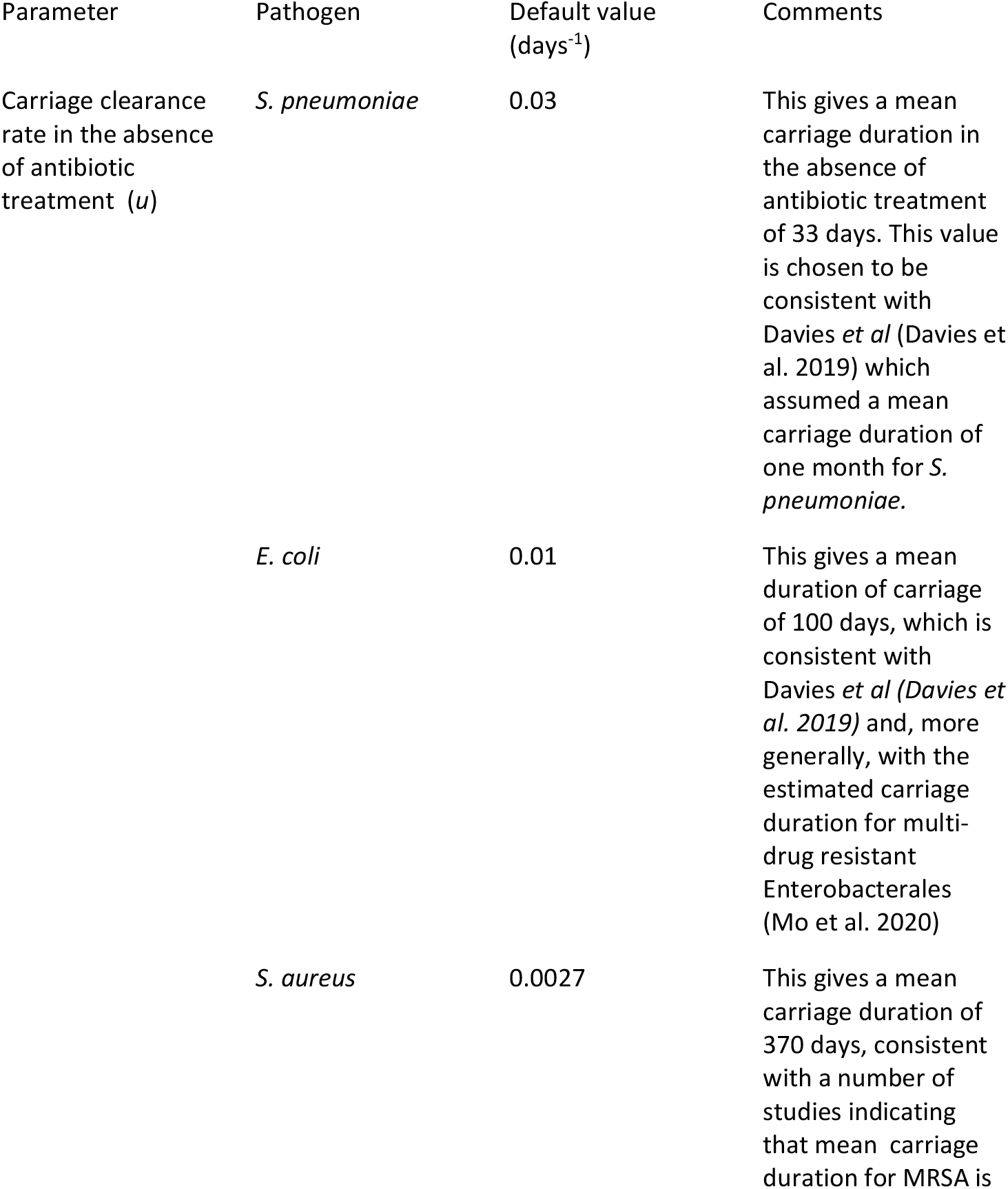
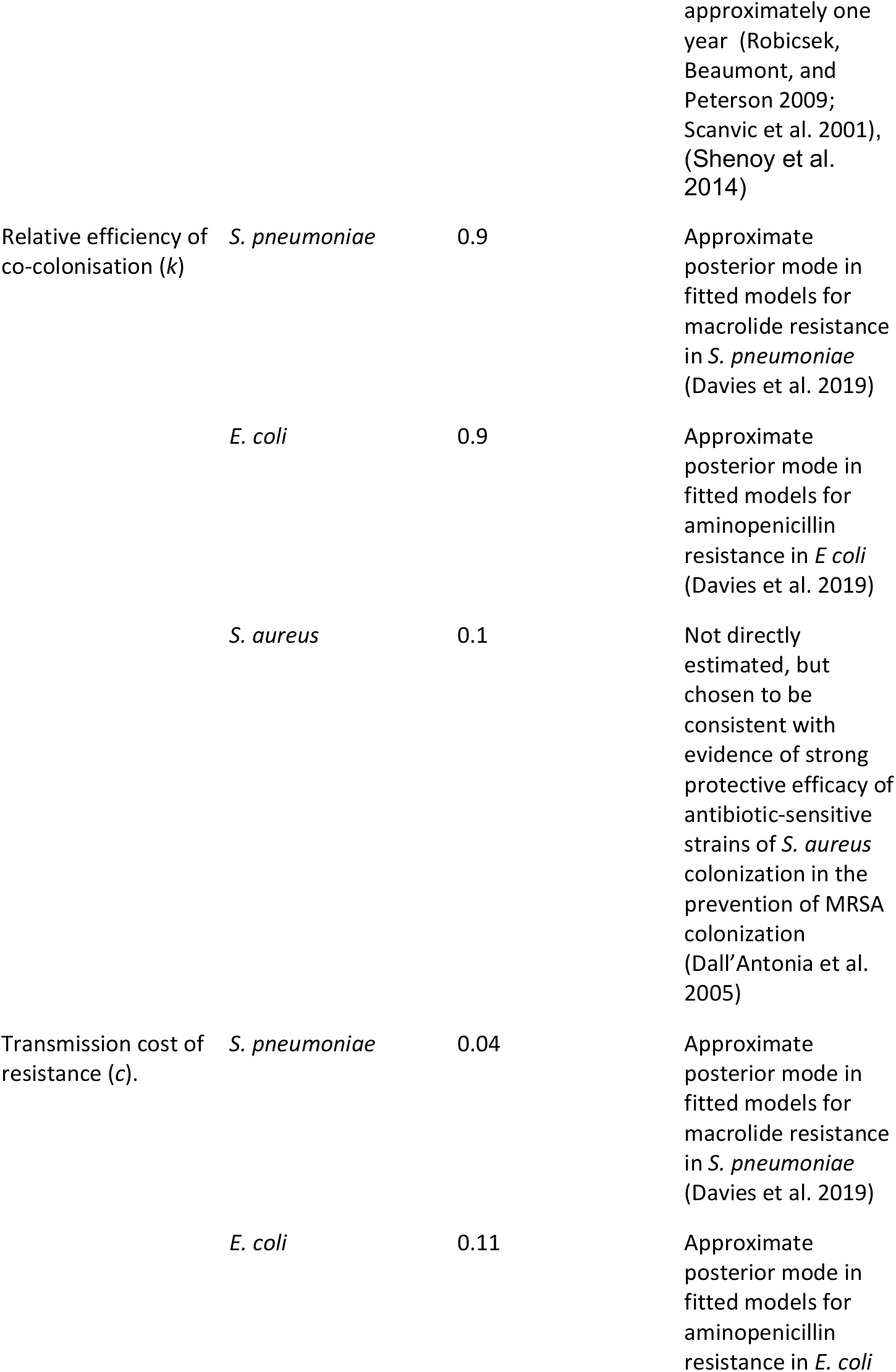
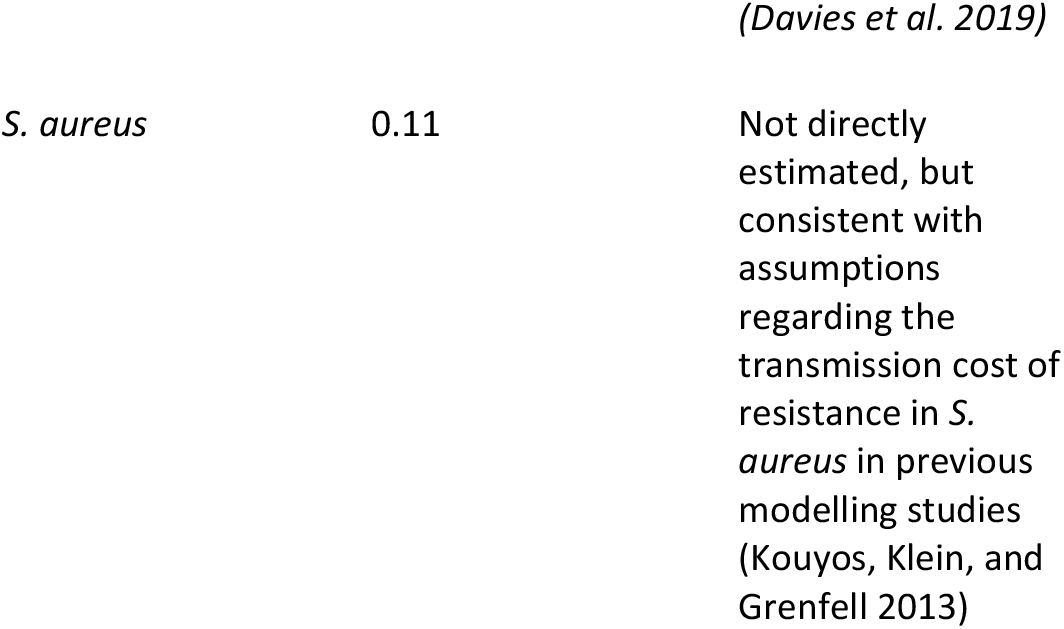
Default values for model parameters which are assumed not to vary by WHO region.

Health benefits of the intervention are measured in terms of disability adjusted life years (DALYs) averted due to reduced risk of sepsis or death in the A-PLUS trial. We used estimates from Patterson *et al. (Patterson et al. 2025)* who used disability weights of 0.0169 for maternal sepsis. The A-PLUS trial did not find any evidence that the intervention led to reduced maternal mortality (relative risk 1.23; 95% CI, 0.51 to 2.97) or to health benefits in infants, so all assumed health benefits were linked to reductions in maternal sepsis.

Adverse effects of the intervention due to DALYs resulting from increased antimicrobial resistance were calculated by taking, for each WHO region, the population weighted average of national estimates of DALYs attributable to macrolide resistance in *S. aureus* and *S. pneumoniae (GBD 2021 Antimicrobial Resistance Collaborators 2024)*, and, for each pathogen, multiplying by 1+ x/100 where x is the percentage increase in resistant infections resulting from the intervention, as determined by the model.

## Results

Predicted changes in macrolide resistance varied substantially according to the model used. The smallest increases in resistance were consistently seen with Model 3 (which accounts for interactions between maternity units and the wider population and includes different age groups) while the largest increases in resistance were seen with Model 1 (which does not include age groups or different settings). Model 2 (which explicitly accounts for maternity units but does not include distinct age groups) showed behaviour that was intermediate between models 1 and 3. For example, in the WHO Africa region, Model 3 predicted that a policy of universal azithromycin prophylaxis during labour would result in an increase in resistance from 33% of *S. aureus* infections to 39% (Table 5), while with models 2 and 1 the increase was to 58% and 78% respectively. There were also consistent differences between regions, reflecting differences in birth rates (Table 2) and hence differences in proportional changes in macrolide usage (Table 3), different baseline levels of resistance and macrolide usage (Tables 3-5), and regional differences in the relationship between antibiotic use and resistance. For all three pathogens considered, the largest increases in resistance caused by the policy occurred in the WHO Africa region, the region with both the highest birth rate and the lowest estimated baseline rate of macrolide usage (Tables 2 and 3). Here Model 3 predicted 16% and 6% absolute increases in macrolide resistance in *S. pneumoniae* and *S. aureus* respectively. The lowest absolute increases in resistance occurred in the Europe and Western Pacific regions (the regions with the lowest per capita birth rates), where Model 3 predicted 2% and 3% absolute increases in macrolide resistance in *S. pneumoniae* and *S. aureus* respectively (Tables 4 and 5).

**Table 2.** Mean births per person per year in 2020.

| WHO region | Mean births per person /year (weighted by population size) |
| --- | --- |
| Africa | 0.034 |
| Americas | 0.013 |
| Eastern Mediterranean | 0.025 |
| Europe | 0.011 |
| South-East Asia | 0.017 |
| Western Pacific | 0.009 |

**Table 3.**
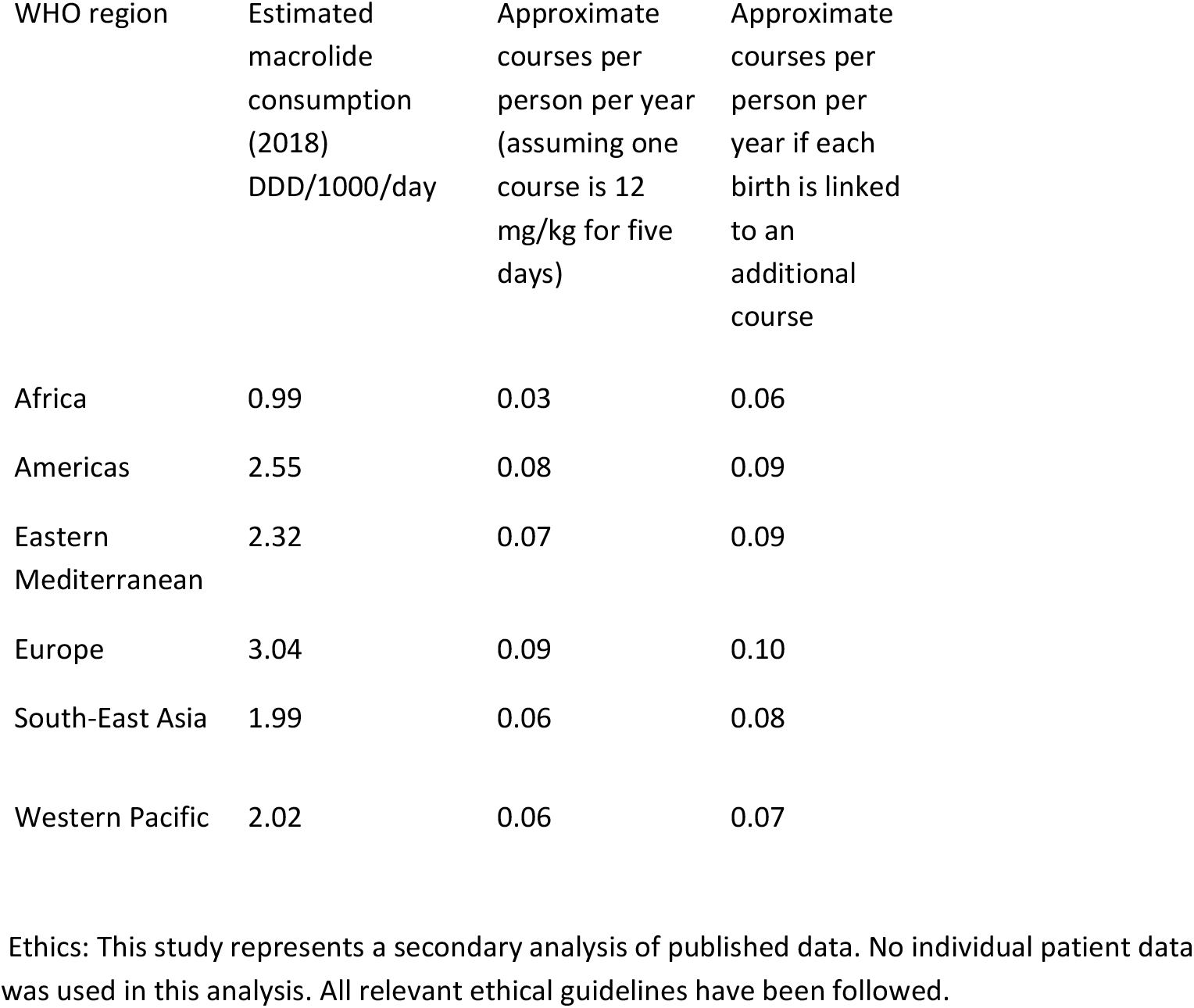
Estimated macrolide use under baseline assumptions and under the modelled scenario where each birth is linked to an additional course.

**Table 4.** Predicted impact on resistance and DALYs due to macrolide resistance increase in *S. pneumoniae* caused by the introduction of azithromycin prophylaxis during labour

| WHO region | Estimated baseline proportion resistant | Predicted proportion resistant with 100% azithromycin prophylaxis | Estimated DALYs that would result from increase in resistance | DALYs per million population |
| --- | --- | --- | --- | --- |
| Africa | 0.38 | Model 1: 0.73 | Model 1: 178,000 | Model 1: 155 |
|  |  | Model 2: 0.59 | Model 2: 108,000 | Model 2: 94 |
|  |  | Model 3: 0.54 | Model 3: 82,000 | Model 3: 72 |
| Americas | 0.38 | Model 1: 0.50 | Model 1: 15,000 | Model 1: 14 |
|  |  | Model 2: 0.45 | Model 2: 9,000 | Model 2: 8 |
|  |  | Model 3: 0.42 | Model 3: 5,000 | Model 3: 5 |
| Eastern Mediterranean | 0.44 | Model 1: 0.64 | Model 1: 32,000 | Model 1: 42 |
|  |  | Model 2: 0.56 | Model 2: 20,000 | Model 2: 26 |
|  |  | Model 3: 0.51 | Model 3: 12,000 | Model 3: 16 |
| Europe | 0.25 | Model 1: 0.35 | Model 1: 11,000 | Model 1: 12 |
|  |  | Model 2: 0.29 | Model 2: 4,000 | Model 2: 4 |
|  |  | Model 3: 0.27 | Model 3: 2,000 | Model 3: 2 |
| South-East Asia | 0.50 | Model 1: 0.61 | Model 1: 60,000 | Model 1: 30 |
|  |  | Model 2: 0.58 | Model 2: 43,000 | Model 2: 21 |
|  |  | Model 3: 0.55 | Model 3: 25,000 | Model 3: 12 |
| Western Pacific | 0.77 | Model 1: 0.80 | Model 1: 8,000 | Model 1: 4 |
|  |  | Model 2: 0.79 | Model 2: 6,000 | Model 2: 3 |
|  |  | Model 3: 0.79 | Model 3: 6,000 | Model 3: 3 |

**Table 5.**
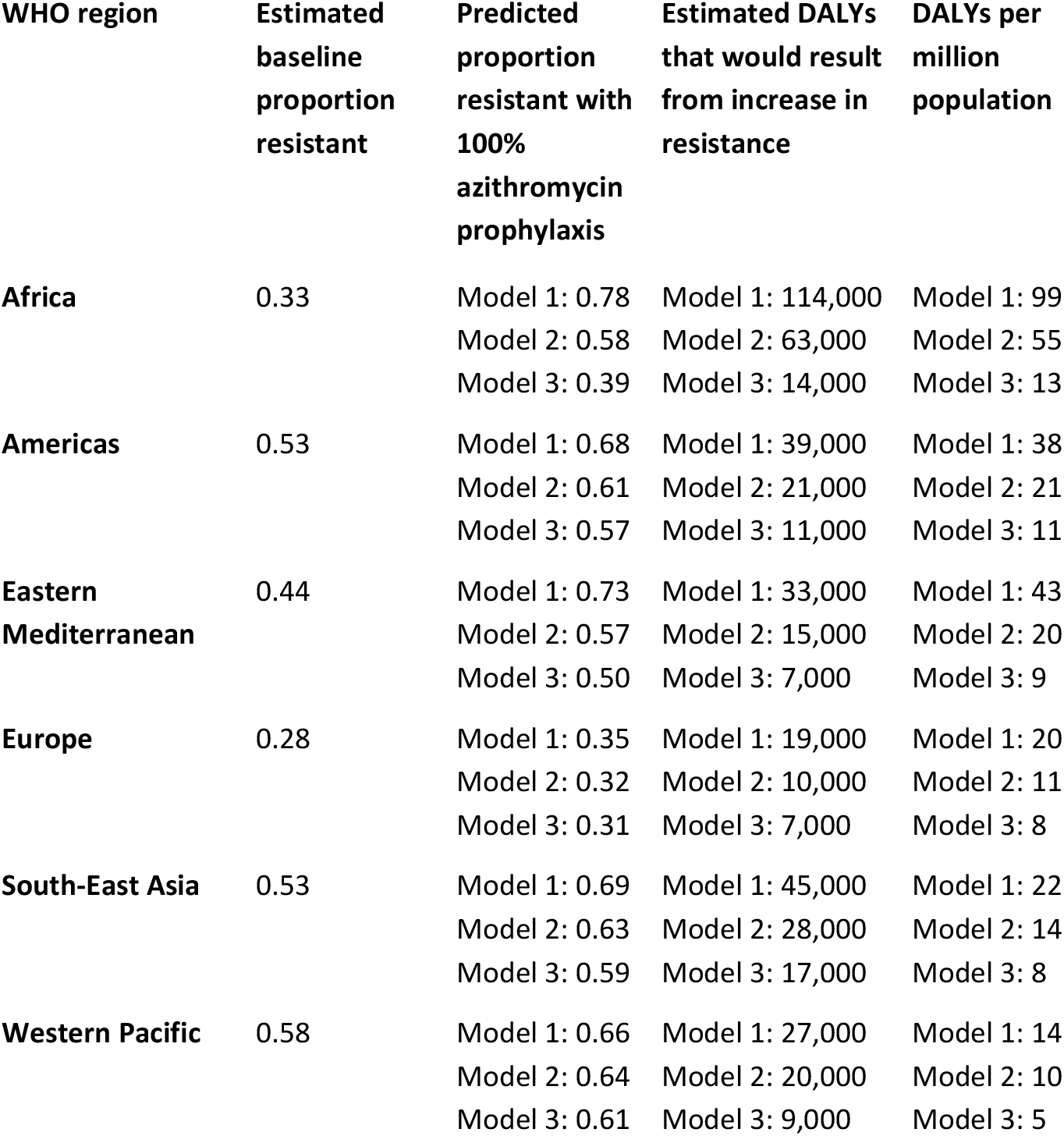
Predicted impact on resistance and DALYs due to macrolide resistance increase in *S. aureus* caused by the introduction of azithromycin prophylaxis during labour

**Table 6.** Predicted impact on macrolide resistance in *E. coli* caused by the introduction of azithromycin prophylaxis during labour under different assumptions about baseline resistance.

|  | Assumed<br>baseline<br>proportion<br>resistant | Predicted proportion resistant with<br>azithromycin prophylaxis |  |  |
| --- | --- | --- | --- | --- |
|  |  | Model 1 | Model 2 | Model 3 |
| Africa | 0.10 | 0.61 | 0.40 | 0.26 |
| Africa | 0.20 | 0.65 | 0.50 | 0.38 |
| Africa | 0.30 | 0.69 | 0.59 | 0.47 |
| Americas | 0.10 | 0.26 | 0.17 | 0.14 |
| Americas | 0.20 | 0.35 | 0.27 | 0.25 |
| Americas | 0.30 | 0.43 | 0.37 | 0.35 |
| Eastern<br>Mediterranean | 0.10 | 0.38 | 0.21 | 0.16 |
| Eastern<br>Mediterranean | 0.20 | 0.44 | 0.33 | 0.27 |
| Eastern<br>Mediterranean | 0.30 | 0.52 | 0.43 | 0.37 |
| Europe | 0.10 | 0.22 | 0.14 | 0.12 |
| Europe | 0.20 | 0.30 | 0.24 | 0.23 |
| Europe | 0.30 | 0.39 | 0.32 | 0.34 |
| South-East Asia | 0.10 | 0.30 | 0.20 | 0.17 |
| South-East Asia | 0.20 | 0.37 | 0.31 | 0.27 |
| South-East Asia | 0.30 | 0.45 | 0.41 | 0.36 |
| Western Pacific | 0.10 | 0.21 | 0.16 | 0.15 |
| Western Pacific | 0.20 | 0.30 | 0.27 | 0.25 |
| Western Pacific | 0.30 | 0.38 | 0.37 | 0.34 |

The adverse health impact of the intervention due to increased resistance in *S. pneumoniae* caused by the intervention was estimated to be substantially higher in the WHO Africa region than in other regions (72 DALYs per million with Model 3, compared with the next highest impact of 16 DALYs per million with Model 3 in the Eastern Mediterranean region). For *S. aureus* the highest adverse impact due to resistance was also predicted to occur in the WHO Africa region (13 DALYs per million with Model 3) though in this case there was substantially less variation between different WHO regions.

These potential adverse impacts of the intervention due to selection for resistance are to be contrasted with potential benefits of the intervention resulting from reductions in maternal sepsis. The A-PLUS trial found no evidence of health benefits in South America, but found that the intervention was linked to a large reduction in maternal sepsis in Africa (RR 0.47 95% CI [0.36, 0.61]) and smaller reduction in Asia (RR 0.88 [0.70, 1.10]) (Tita et al. 2023). This led to estimates of 27.7 (95% CI [23.8, 31.8]) DALYs averted by the intervention per 100,000 pregnancies in Africa and 4.2 (95% CI [-3.3, 10.7]) in Asia (Patterson et al. 2025). Based on UN data there were 38.59 million live births in the WHO Africa region (AFR) in 2020, 34.31 million in the WHO South-East Asia region (SEAR), and 18.15 million in the Western Pacific region (WPR). Using the estimated DALYs averted per pregnancy from Patterson *et al* (with the estimate for Africa applied to the AFR region and the estimate for Asia applied to the SEAR and WPR regions, and approximating the annual number of pregnancies leading to labour in each region by the UN live birth data) gives annual estimates of DALYs averted of 10,689 (95% CI [9,184, 12,272]) in AFR, 1,441 (95% CI [-1,132, 3,671]) in SEAR and 762 (95% CI [-599, 1942] in WPR.

The results of this analysis indicate that for all three WHO regions and with all model scenarios considered, the indirect negative health impacts of the intervention (measured in DALYs due to increased macrolide resistance in *S. aureus* and *S. pneumoniae*) were substantially greater than the positive health impacts, the former being 27 to 73 times higher than the latter when using model 1, and 9 to 29 times higher when using model 3.

A threshold analysis indicated that, under the above assumptions, if the intervention resulted in an absolute increase in population-wide macrolide resistance in *S. aureus* and *S. pneumoniae* in AFR of 1.4% or higher, or absolute increases of 0.17% or higher in SEAR or 0.13% or higher in WPR then the negative health impacts (measured in DALYs) would outweigh the health benefits of the intervention. These thresholds are below the expected increases in resistance in all models considered.

## Discussion

The model results suggest that unless a policy of routine azithromycin prophylaxis for pregnant women in labour planning a vaginal birth has additional health benefits beyond the demonstrated reduction in maternal sepsis (for example, reduction in maternal mortality) it is plausible that the negative health impacts due to selection for resistance would outweigh the positive health benefits due to reduced sepsis.

The analysis indicates that the per capita adverse health impacts of the policy are likely to be largest in the WHO Africa region. This is, in part, a consequence of the higher birth rate in this region which means that routine azithromycin prophylaxis during labour would lead to a greater per capita increase in azithromycin usage than in other regions. An additional factor is that, compared to other regions, there is evidence of a higher elasticity in the relationship between antibiotic use and resistance in Africa; that is, for a given increase in antibiotic use we expect to see a higher change in resistance. This can be seen in the high estimated rates of estimated macrolide resistance in *S. aureus* and *S. pneumoniae* despite low estimated rates of macrolide consumption in Africa. For example, estimated rates of macrolide resistance in both *S. aureus* and *S. pneumoniae* are higher in Africa compared to Europe (33% versus 28% and 38% versus 25% respectively) despite Europe having an estimated per capita macrolide consumption that is three times higher than that in Africa. While it is possible that this might be partly explained by distortions in clinical data collection leading to biased estimates (there are important limitations with both resistance data and antibiotic use estimates which we discuss later), observational studies which have prospectively collected bacterial carriage data and antibiotic usage data in sub-Saharan Africa have repeatedly found high levels of antimicrobial resistance despite low levels of antibiotic use (Lewis et al. 2022, 2019; Heinemann et al. 2023).

Direct population-level evaluations of the impact on antimicrobial resistance of the introduction of routine azithromycin prophylaxis during labour are lacking. There are, however, a number of tangential studies which provide important background for the modelling results. In a population-based cross-sectional pneumococcal carriage study of children in Malawi which followed up communities who took part in a cluster randomised placebo controlled trial of an MDA programme in which pre-school children received twice-yearly azithromycin or placebo for 2 years, Kalizang’oma *et al* found that 3.5 years after cessation of the MDA programme, macrolide resistance in *S. pneumoniae* had increased from a baseline of 22% to 32% (Kalizang’oma et al. 2025). Similar increases were seen in nearby communities who were in the control group, but no such increase in macrolide resistance was seen at a more distant control site where MDA had not been implemented. This study provides evidence that the azithromycin use in the MDA intervention was the cause of substantial and persistent population-level increases in macrolide resistance. An earlier study in Niger which followed up children in the same trial 6 months after the last administration of azithromycin also found evidence of substantial increases in determinants of macrolide resistance in the intestinal flora, with an absolute increase of over 20% (Doan et al. 2019).

Mass drug administration of azithromycin is also a key component in trachoma control programmes, and a systematic review assessed the evidence that this intervention was linked to increases in macrolide resistance in *S. pneumoniae, S. aureus, E. coli* as well as *Chlamydia trachomatis (O’Brien et al. 2019)*. Overall, the authors concluded, there was consistent evidence that the intervention led to substantial increases in macrolide resistance in *S. pneumoniae* in communities receiving the intervention though these increases were reversed when MDA stopped. There was also more limited evidence that the intervention led to an increase in macrolide resistance in *S. aureu*s and *E. coli*. There was no evidence of clinically significant azithromycin resistance in *C. trachomatis*.

A Cochrane review has also summarised evidence of increased risk of resistance to azithromycin in *S. pneumoniae, S. aureus*, and *E. coli* in communities treated with azithromycin as well as evidence of increased cross-resistance to tetracycline and clindamycin (Evans et al. 2019). When azithromycin resistance was present at baseline, absolute increases in percentages of children carrying resistant strains were typically between about 20% and 40%.

The impact of oral azithromycin taken during labour has been studied at an individual level: Roca et al. reported large short-term (28 day post-treatment) increases in the prevalence of azithromycin resistance in *S. aureus* and *S. pneumoniae* recovered from nasopharyngeal swabs from treated mothers in The Gambia (Roca et al. 2016). This study also found short-term increases in azithromycin resistance in *S. aureus* recovered from nasopharyngeal swabs from infants of treated mothers. However, 12 months after delivery, the prevalence of azithromycin resistance in *S. aureus* and *S. pneumoniae* had waned to baseline levels (Bojang et al. 2018).

Considered together, these results provide evidence that while at an individual level elevated levels of carriage of resistance strains may be relatively short-lived, community-wide increases in the use of azithromycin would be expected to lead to population-level increases in azithromycin resistance in *S. aureu*s, *S. pneumoniae* and *E. coli*.

There are a number of important limitations to our study that need to be highlighted. First, we model the increase in macrolide use as an additional course, as our models follow Davies et al (Davies et al. 2019) in relating courses of antibiotic use per person per year to changes in resistance, rather than other measures of usage (e.g. defined daily doses or weight). This represents an implicit assumption that the number of distinct episodes of antibiotic exposure that an individual experiences is a relevant quantity for understanding resistance selection. While we believe there are theoretical reasons supporting this approach, we recognise that alternative approaches are possible.

A second limitation that we do not consider cross-resistance in the models.There is evidence that this is potentially important when considering selection of resistance through increased use of azithromycin and its inclusion would be expected to increase the adverse health effects of the intervention (Kalizang’oma et al. 2025; Evans et al. 2019)

A third limitation is that, in this initial work, we do not explicitly model different *S. pneumoniae* serotypes or explicitly account for the role of adaptive immunity in shaping *S. pneumoniae* carriage dynamics. There would be value in extending the analysis to include the serotype specific model (and extensions of this) described by Davies et al. (Davies et al. 2019). More generally, as with all models, a number of simplifying assumptions are made and while our baseline model was better able to explain the relationship between antibiotic use and resistance than simpler models when applied to European data, there would be value in a more thorough assessment of the predictive capability of the models used. We note, however, that the DALYs estimated to result from increased resistance in a single pathogen, *S. aureus* (where such immunity-mediated interactions are not relevant) already considerably exceed the DALYs estimated to be averted by the intervention.

A further limitation is that our model does not explicitly account for reductions in antibiotic use that would be expected to result from the reduction in sepsis caused by the intervention, and we note that Patterson estimated that approximately 1800 courses of antibiotics could be averted by the intervention per 100,00 pregnancies (Patterson et al. 2025).

Another important area for future work is quantifying uncertainty associated with our findings. However, the very large discrepancy between the health benefits of the intervention estimated by Patterson *et al* and the adverse health impacts estimated here mean that it is very unlikely that this additional analysis will alter the overall conclusions in any important way.

When calibrating models to data, an additional limitation which would apply to all modelling approaches is the lack of validated antibiotic use data outside high income countries. Existing estimates of antibiotic usage rest primarily on proprietary antibiotic sales data which may fail to capture informal selling of antibiotics. Indeed, in the few cases when different sources of antibiotic use data are available in lower and middle income countries, large discrepancies have been observed (A. J. Browne 2021). Similarly, estimates of macrolide-resistance in *S. aureus* and *S. pneumoniae* are dependent on imperfect data collected for clinical rather than surveillance purposes and may also be subject to important biases, particularly in settings where susceptibility testing is predominantly performed when patients do no respond to initial treatment (Lim et al. 2021). Estimates of DALYs attributable to resistance are, likewise, vulnerable to a number of important biases as they are, to a large extent, dependent on estimated relative risks of mortality associated with antimicrobial resistance and only a small percentage of these constituent estimates can be considered to be at low risk of bias(Hassoun-Kheir et al. 2024). A further issue is that the estimated DALYs attributable to resistance are based on a comparison of outcomes between drug-resistant and drug-sensitive infections, implicitly assuming that increases in resistant infections will be matched by concomitant decreases in susceptible infections with the same bacterial species (Marlieke E. A. de Kraker and Lipsitch 2022). While this seems plausible for *S. pneumoniae*, this is not generally true and for at least some bug-drug combinations there is clear evidence that increases in the resistant phenotype are not accompanied by decreases in the susceptible phenotype(Mostofsky, Lipsitch, and Regev-Yochay 2011; M. E. A. de Kraker et al. 2013; Ammerlaan et al. 2013). If this is also the case for macrolide resistance in *S. aureus*, then the estimates for attributable DALYs used would likely represent substantial underestimates of the true values, meaning that we would be underestimating the indirect negative health impacts of the intervention.

It is important to recognise that the above results are predicated on the assumption that the intervention has no effect on maternal mortality. While the A-PLUS trial did not demonstrate that the intervention reduced maternal mortality, it did find a large reduction in maternal sepsis in Africa, with some evidence of a smaller reduction in maternal sepsis in Asia (though, in the latter case, the 95% confidence interval was also compatible with either no effect or a small increase in maternal sepsis). If the reduction in sepsis were to be the cause of even a modest reduction in mortality we might reach substantially different conclusions: for example, if in 10% of the sepsis cases prevented the intervention also prevented one maternal death, then the DALYs averted by the intervention would be two orders of magnitude higher than those estimated by Patterson et al. (Patterson et al. 2025) and likely to considerably outweigh the DALY increase due to increased resistance resulting from the intervention. However, if we assume no impact on maternal mortality and consider only the health benefits of the intervention included by Patterson et al. then under all model scenarios considered in our analysis the negative health impacts of the intervention would be expected to considerably outweigh the health benefits.

## Data Availability

All data produced in the present work are contained in the manuscript

## Transparency Declaration

None to declare.

## Appendix 1

## Model 1

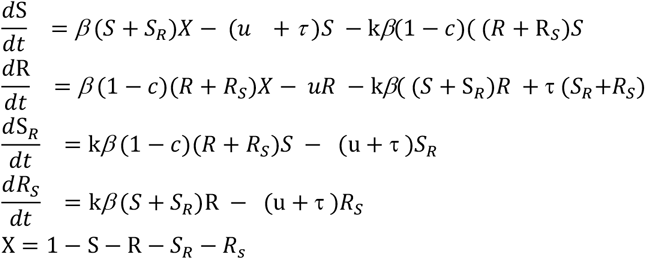

See figure 1 for variable names and Table 1 for parameter names for parameters which are assumed not to vary with WHO region. Parameters which do vary with WHO region are treatment induced clearance rate (***τ***) and the transmission parameter (β)..

## Model 2

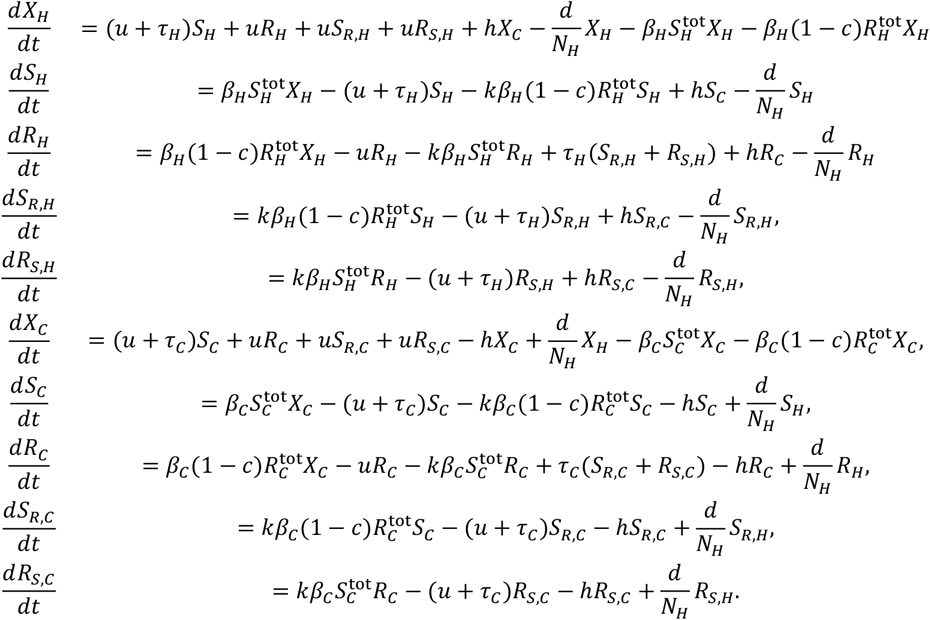

Where

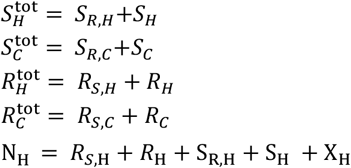

Variable name are as described in model 1, but with *C* and *H* subscripts corresponding to hosts in the community and hospital respectively. Parameters are as for figure 2. The new parameter *h*, represents the hospital admission rate.

## Model 3

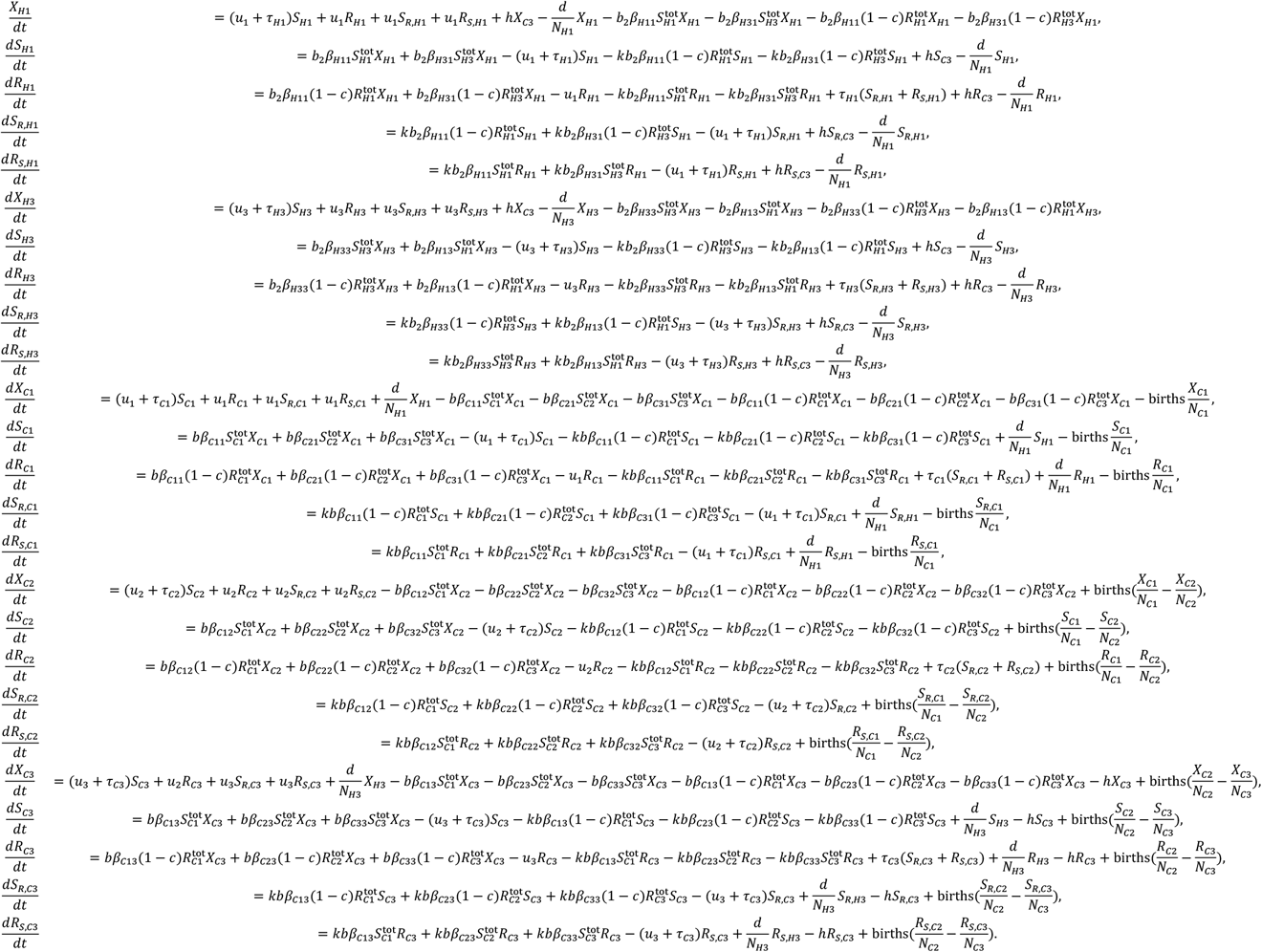

Where

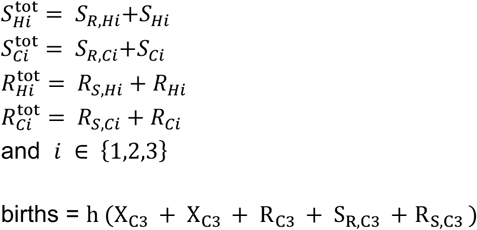

The terms N_*Hi*_ and N_*Ci*_ represent the total population sizes in age group i in the hospital and community respectively, and b represents a scaling factor for transmission in the community.

